# Out of hospital cardiac arrests in the Gulf Region: a scoping review

**DOI:** 10.1101/19014118

**Authors:** Alan M. Batt, Chelsea Lanos, Shannon Delport, Dalal Al-Hasan, Shane Knox, Assim Alhmoudi, Megan Anderson, Saleh Fares, Fergal H. Cummins

## Abstract

**Background:** Out-of-hospital cardiac arrest (OHCA) is a major cause of mortality worldwide. Recent studies demonstrated low survival rates in Middle Eastern countries. Anecdotally there are unique demographic, cultural and logistical challenges in this region. However, there remains a paucity of data published on OHCA in the Middle East. In order to address OHCA in a meaningful manner in the region, we first need to quantify the issue.

**Methods:** We conducted a scoping review of published and grey literature on OHCA in the Gulf Cooperative Council region that utilised Arksey and O’Malley’s framework. Electronic databases and grey literature sources were identified and searched. Subject matter experts in the region were consulted. All types of studies in English and Arabic were included.

**Results:** A total of 24 studies were included from Saudi Arabia, the UAE, Oman, Kuwait, and Qatar. No literature was identified from the state of Bahrain. OHCA victims in the region are younger, predominantly male, and more co-morbid than other international studies. We observed low Emergency Medical Service utilisation, low bystander cardiopulmonary resuscitation, return of spontaneous circulation, and survival to discharge rates across the region. There are differences in characteristics of OHCA among ethnic groups.

**Discussion and conclusions:** We identified unique characteristics associated with OHCA in the region, variances in processes and outcomes, and a lack of coordinated effort to research and address OHCA. We recommend creating lead agencies responsible for coordinating and developing strategies such as community response, public education, and reporting databases.

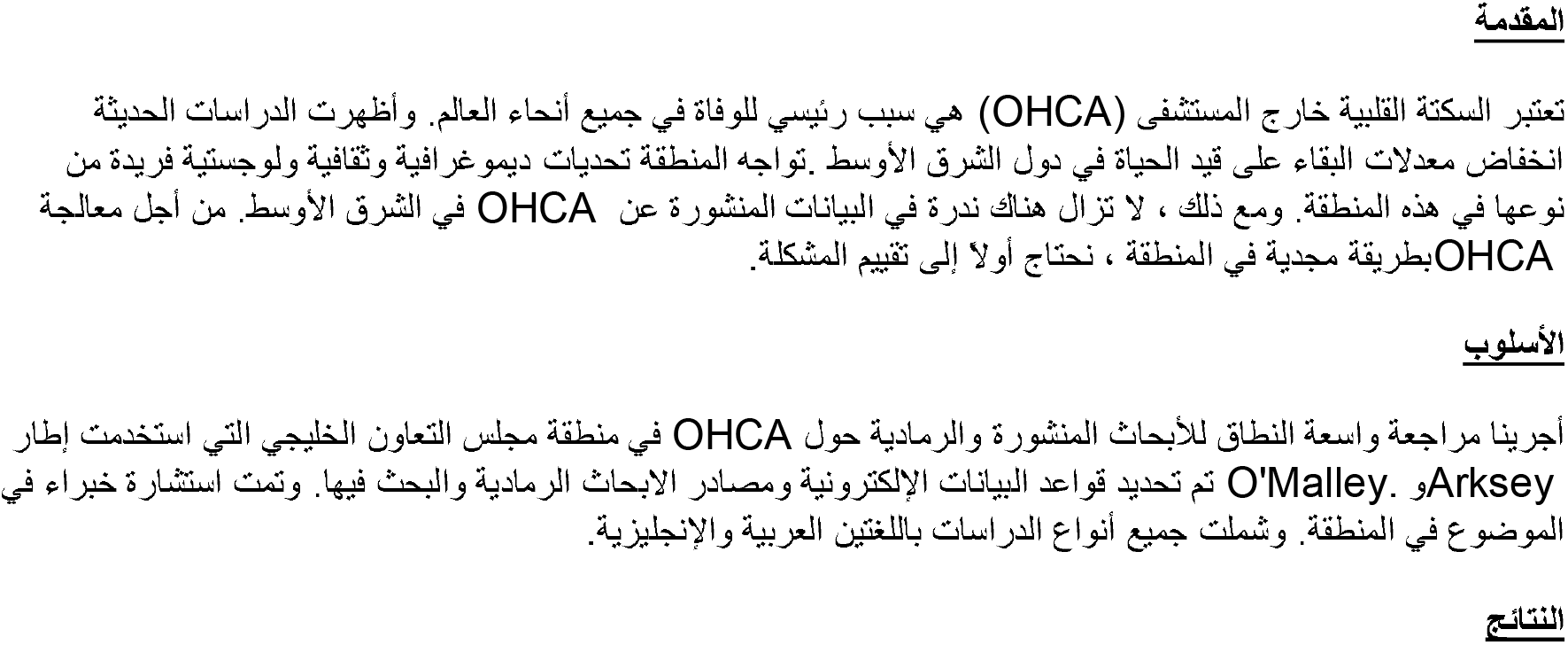

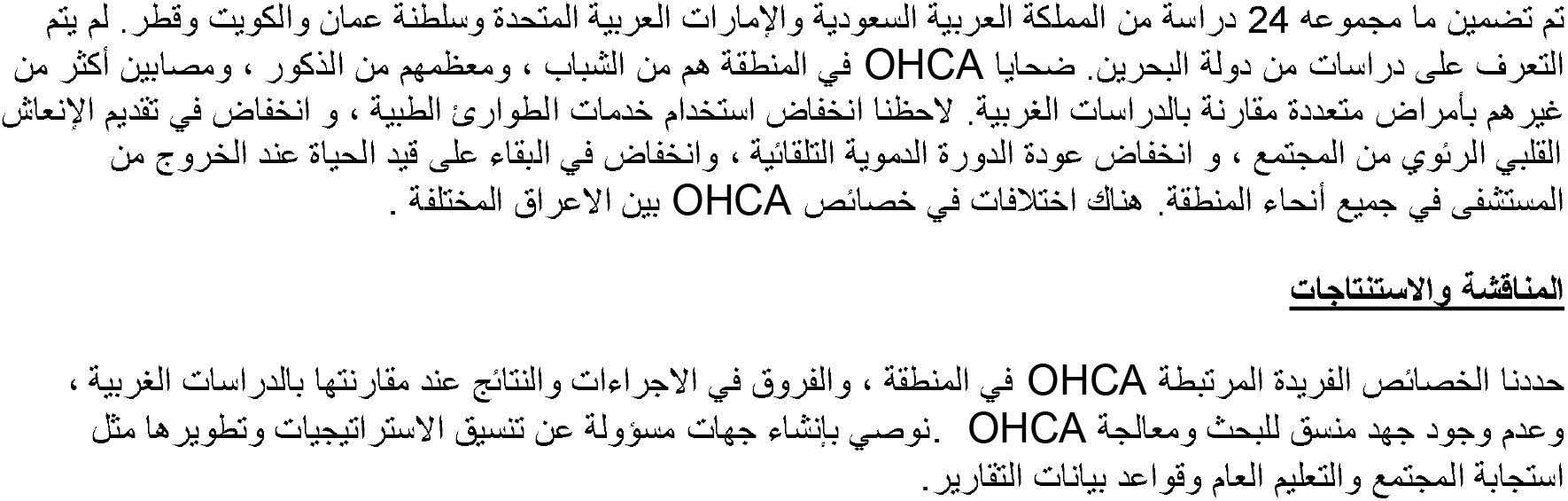

## Introduction

Out-of-hospital cardiac arrest (OHCA) is a major cause of mortality worldwide, with variable survival reported across countries and systems. In particular, Middle-Eastern and South Asian countries report low survival rates (1). Reported reasons for low OHCA survival in these contexts include unique demographic, cultural, and logistical challenges related to the management of OHCA. Such contexts may also present a challenge when we attempt to implement evidence produced for the most part in vastly different systems. Characteristics and outcomes of OHCA in Middle Eastern countries remains poorly researched, compounding these challenges. The resulting lack of knowledge prevents us from understanding the issue, and thereby prevents us from improving the issue. Thus, to meaningfully address OHCA in the Middle East, we first need to understand the issue within its context.

Within the Middle East, the Cooperation Council for the Arab States of the Gulf, also known as the Gulf Cooperation Council (GCC, 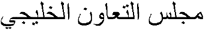) was established in 1981, and comprises the governments of the states of the Kingdom of Bahrain, Kuwait, the Sultanate of Oman, Qatar, the Kingdom of Saudi Arabia, and the United Arab Emirates (UAE). The countries in the GCC are predominantly Islamic, and they share a common cultural and historical background (2). Another shared trait is their economic reliance on expatriate workforces (3), estimated at 48% of the total regional population of 53 million (4,5). Within this regional population life expectancy has risen dramatically from 42-61 years since the 1960s to 74-80 years in 2018 (5). However, increased population numbers, the healthcare needs of different demographics and increased life expectancy have led to a rise in healthcare demands. This rise is predominantly due to high rates of non-communicable diseases such as diabetes, cardiovascular disease, and obesity, attributable to unhealthy lifestyles and a general lack of coordinated public health programmes (4,6). In order to reduce the risk associated with such morbidity, the majority of GCC member countries have initiated public health strategies which include the recording and reporting of non-communicable diseases

Sasser et al. first reported on out-of-hospital emergency care in Abu Dhabi in 2004 (7), when emergency medicine in the GCC was rudimentary at best. They observed no real medical control, no treatment policies, a lack of training and education, and no quality assurance policies, which echoed the situation across the region. However, emergency medicine and prehospital care have developed across the GCC in the last 15 years. Reporting of OHCA rates, response, and outcomes are also considered benchmarks of emergency medical response readiness. As a result, we elected to conduct a review of OHCA studies in the GCC in order to collectively summarise their characteristics and properties, as well as to identify any gaps and areas for further research.

## Methods and analysis

We conducted a scoping review, which enabled us to identify, map and present an overview of a heterogeneous body of literature (8,9). We were interested in exploring the topic of OHCA within the region, and comparisons to other international systems are not intended to compare the effectiveness or appropriateness of either system. We deemed a scoping review to be appropriate given the predicted lack of published data (and potential reliance on grey-literature sources), the anticipated heterogeneity among studies, and the expected variability in reporting. We performed a systematic literature search for studies that investigated OHCA in the GCC region. We utilised Arksey and O’Malley’s five-stage framework which included [1] identifying the research question, [2] identifying relevant studies, [3] refining the study selection criteria, [4] collecting relevant data from each article, and [5] collating, summarizing, reporting, and interpreting the results. We also implemented a recommended additional stage, [6] consultation with experts. We reported our process according to the PRISMA Extension for Scoping Reviews (10).

### 1. Identify the research questions

1. What are the characteristics of out-of-hospital cardiac arrest cases in the GCC region?
2. How do these studies differ from other international studies, evidence from which are predominantly used to inform resuscitation guidelines?
3. What insights can be gained from these findings in order to better inform policy and practice in the GCC region?

#### Ethics and dissemination

This is a scoping review of completed studies and hence no ethics approval was required.

### 2. Identify relevant studies

Systematic search

An information scientist (MA) developed the search strategy. We used this to search the electronic databases MEDLINE, CINAHL, Web of Science and EMBASE from 1990 until June 2019. The search strategy was modified accordingly for each database. Relevant nonindexed journals were identified and searched, and we performed multiple Google web and Google Scholar searches. We searched the grey literature with guidance from the Grey Matters checklist (11). References of included studies were scanned for relevant articles. See Appendix I for search terms and a list of non-indexed sources.

### 3. Select the studies

#### Eligibility criteria

Studies in English and Arabic were eligible for inclusion if they reported OHCA in the GCC. We excluded studies that were not related to OHCA (e.g. in-hospital cardiac arrest), case-reports, studies that reported only on traumatic OHCA, studies that were related to cardiopulmonary resuscitation (CPR) training, and those not conducted in GCC countries.

#### Title and abstract screening

Screening and selection comprised of a review of title and abstracts by two reviewers (AB and CL). We resolved disagreements through discussion until consensus was achieved. Where disagreement remained or there was insufficient evidence to make a decision, we included the citation for full text review and subsequent decision.

### 4. Chart the data

To support the full-text review, we developed a standardised data extraction form to organize information, confirm relevance, and to extract study characteristics (See Appendix II) (12). Two reviewers (AB and CL) abstracted general study characteristics such as authorship, year of publication, country, study design, sample size, outcomes measured, and OHCA characteristics. We compiled all data into a single spreadsheet in Microsoft Excel 2013 (Microsoft, Redmond, WA) for analysis.

### 5. Collate, summarize, report and interpret

#### Data summary and synthesis

Due to variations in study designs, terminology used, and reporting conventions, it was necessary to identify common areas for reporting summarised results. We selected a number of demographic, process and outcome measures that reflect key components of effective OHCA response and management. We abstracted and reported the findings for each measure per study where possible, and we report on them under the headings of characteristics, process and outcomes measures.

#### Critical appraisal

In line with the scoping review framework, we did not conduct a critical appraisal (8).

### 6. Consultation (optional stage)

In addition to our own experience and expertise with OHCA in the region, we conferred with a number of OHCA subject matter experts (SMEs) in the GCC, and invited them to contribute additional materials and unpublished studies. Two of these individuals subsequently joined the research team (DA and AA).

## Results

### Search results and study selection

The search yielded 590 citations. We identified an additional 57 citations through searches of grey literature and hand searching. After elimination of duplicates, we screened 284 citations at the title and abstract level. This led to the exclusion of 236 citations. After full-text review of 48 citations, we included 24 full-texts for data extraction and analysis. See Fig 1 for an illustration of these findings using PRISMA Diagram, Appendix III for details of adult OHCA studies, and Appendix IV for details of paediatric OHCA studies.

**Fig 1.**
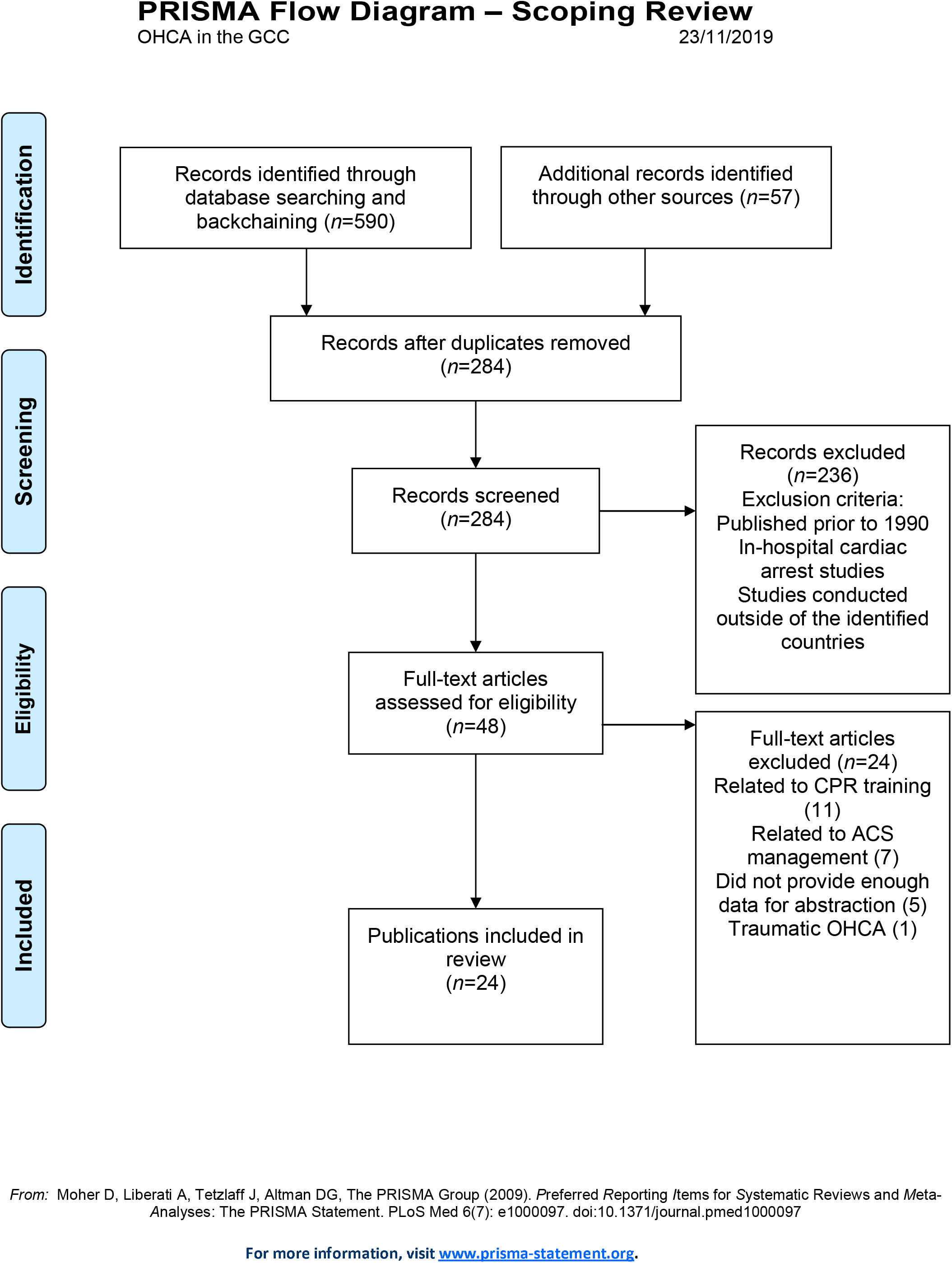
PRISMA diagram of search results.

### Characteristics of included studies

Included studies were published between 1999 and 2019. Studies comprised full-text articles (both published and unpublished manuscripts) (n=16), technical reports or theses (n=3), conference abstracts (n=3), and letters to the editor (n=2). Studies were from the UAE (n=10), Qatar (n=6), Saudi Arabia (n=4), Kuwait (n=3), Oman (n=1), and the region (n=1). We did not identify any literature from the state of Bahrain despite consultation with an SME in the country. A total of 16 studies (across 21 sources) described adult cases or all cases of OHCA. Four studies specifically or independently described paediatric OHCA and we reported these separately in our results; Conroy (1999) (13) is reported as both an adult and a paediatric OHCA study. Where results were published in more than one source (for example a technical report and a peer-reviewed manuscript), we only abstracted the most complete version. Full details of which sources were abstracted is contained in Appendix III.

### Characteristics of adult OHCA studies

Adult OHCA victims in the GCC region were consistently young, predominantly male, and often had underlying co-morbidities. Most OHCA cases occurred at home (ranging from 54% to 85% of all OHCA cases), consistent with international findings (14). See Table 1, Fig 2 and Fig 3 for further characteristics, and Appendix III for full details.

Table 1. Key characteristics, process and outcome measures of abstracted OHCA studies.

*note: Al Hasan 2019c is an unpublished manuscript.

**Fig 2.**
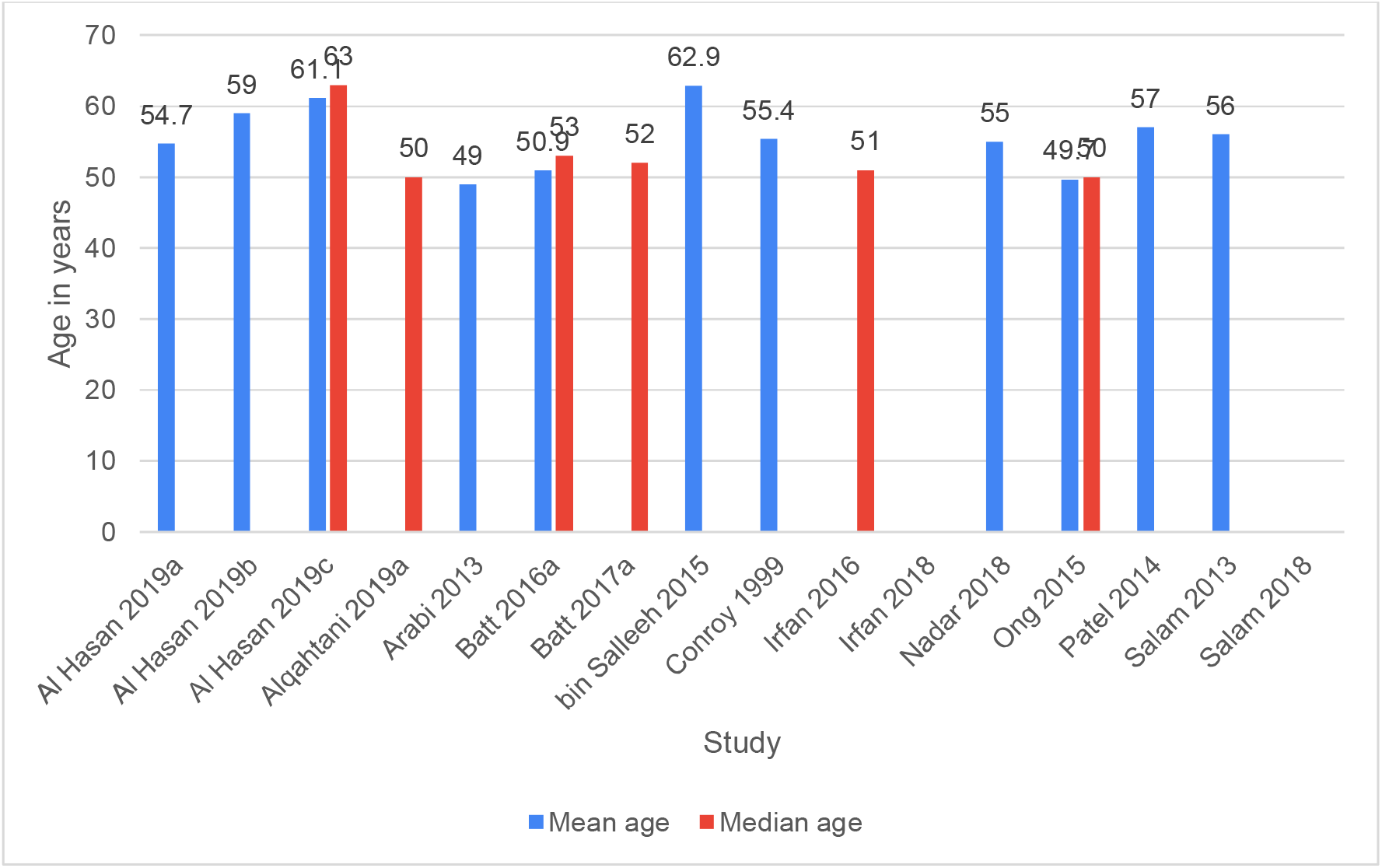
Age profile of included adult OHCA studies.

**Fig 3.**
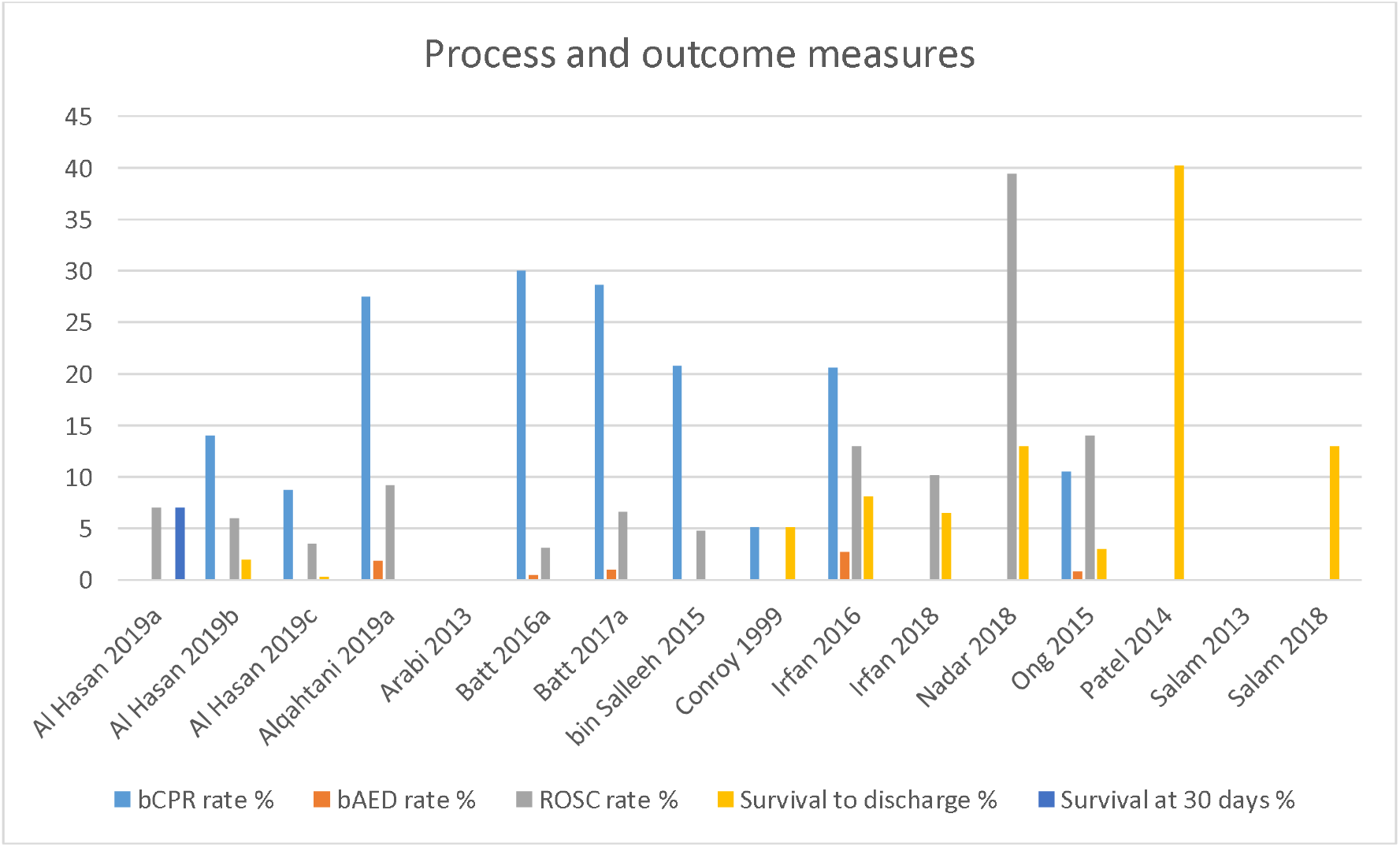
Process and outcome results of included adult OHCA studies.

We observed differences in the characteristics of OHCA among ethnic groups in the region (e.g. Arab, South Asian and North African). For example, compared to GCC national patients, South Asian and North African patients were more likely to smoke, but had lower levels of comorbidities (18,23). Despite this, South Asian and North African patients were generally younger, and accounted for a large percentage of total OHCA cases in a number of studies (17,19,22), possibly attributable to population demographics. Data on specific process and outcome findings for national populations was not well reported across the studies. We abstracted such data where it was available (see Appendix III).

### Process and outcome characteristics

We observed low levels of EMS utilisation, low bystander CPR rates, low return of spontaneous circulation (ROSC) rates, and lower survival to hospital discharge rates across the region when compared to international figures. EMS utilization rates were rarely reported, and ranged from 1.4% (24) to 31% (21). Bystander CPR (bCPR) rates ranged from 5.1% (13) to 30% (19). Bystander AED (bAED) rates were reported at under 2% for any study which reported such data. ROSC rates ranged from 1.6% (16) to 39.4% (24) while most studies reported ROSC rates under 10%. Survival to hospital discharge rates were often not reported, and ranged from 2% (16) to 40.2% (24); however, most studies reported rates under 10%. See Fig 3 for information on process and outcome results.

### Characteristics of paediatric OHCA cases

A small number [4] of paediatric OHCA (POHCA) studies were identified. Despite the limitations associated with such a small number of studies, we report them here nonetheless in order to provide some collective insight into POHCA in the GCC. POHCA victims in the GCC region were predominantly male, and a significant percentage were due to non-medical aetiologies (23.5%-45%), including drowning. Most POHCA cases occurred at home, ranging from 36% (29) to 87% (30) of all POHCA cases. Despite this, and similar to adult OHCA findings, we observed low levels of bystander CPR. See Table 1 and Fig 5 for further characteristics, and Appendix IV for full details.

**Fig 4.**
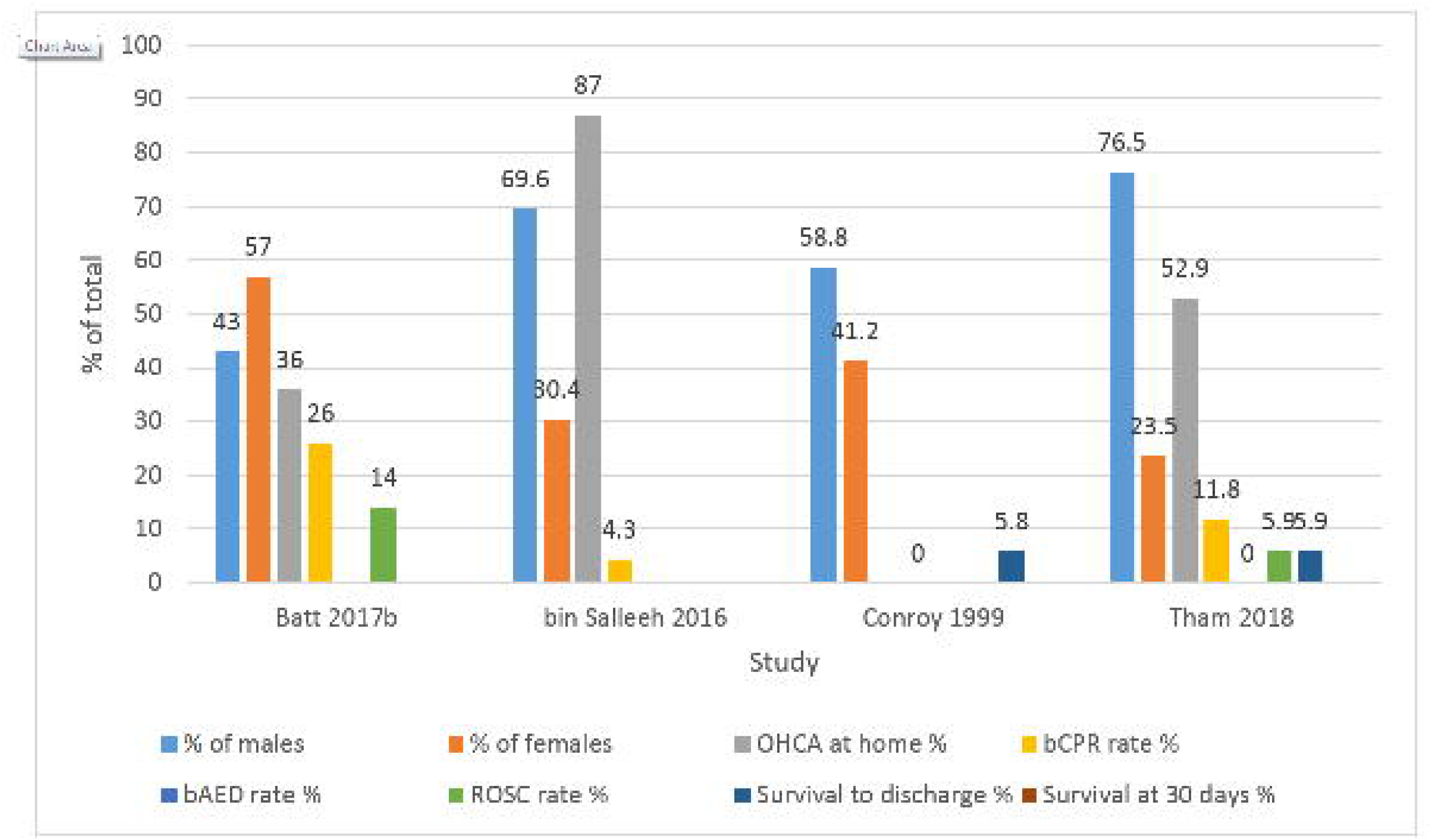
Demographic, process and outcome results for included paediatric OHCA studies.

### Process and outcome characteristics

We observed low bCPR rates, low ROSC rates, and low survival to hospital discharge rates across POHCA studies in the region. Bystander CPR (bCPR) ranged from 0% (13) to 26% (29). No bAED use was reported for any POHCA case in the region. Two studies reported ROSC, 5.9% (31) and 14% (29). Two studies reported a survival to discharge rate of approximately 6% (13,31). Pre-existing medical conditions were reported for POHCA patients in several studies and varied from 23.5% (31) to almost 70% (30). See Fig 5 for information on process and outcome results for POHCA.

## Discussion

The causes of lower OHCA survival in the GCC are anecdotally related to unique demographic, cultural and logistical contexts of the region. Given these challenges, and the lack of synthesized literature, we sought to describe and explore OHCA in the GCC via a scoping review methodology. Included studies reported low out-of-hospital cardiac arrest survival to discharge rates (1-13%) when compared to other international studies [e.g. 1023% reported in Gräsner (32)]. After we examined studies from multiple countries in the GCC region, we suggest several unique characteristics are associated with OHCA in the region; variances exist in both OHCA processes and outcomes; and, despite the mortality associated with OHCA in the region it remains poorly researched, with an observed lack of coordinated effort to improve process and outcomes.

### Unique characteristics of OHCA in the region

OHCA victims in the GCC are younger, predominantly male, and more co-morbid than their counterparts in other international studies (see Appendix III for full details). We suggest a number of reasons for these findings. One proposed cause for the observed increase in cardiac conditions is oil wealth-induced “*obesogenic urbanization”* that has occurred rapidly in the region (33). A number of studies in our review reported high rates of cardiac risk factors including diabetes mellitus, hypertension, hyperlipidaemia, chronic renal failure, and acute coronary syndrome (18,19,23,24,26,28,34). Additionally, the high rate of expatriate populations living in some GCC countries (17,19,22), and their unique lifestyle risk factors (such as high rates of smoking) may have contributed to the observed young age and male dominance of OHCA cases in our review. We also observed differences in OHCA demographics in these populations compared to the national populations. For example, expatriates were normally younger, and more likely to smoke compared to nationals (18,23).

### Variances in pre-hospital resuscitation processes and outcomes

Cardiac arrest cases in public places have generally demonstrated better outcomes in international studies, likely due to an increased rate of witnessed arrests, early activation of EMS and early provision of bystander CPR and AED. Existing evidence acknowledges this portion of care prior to EMS arrival is crucial in OHCA outcomes, i.e. recognising OHCA, calling for help and providing bystander CPR and AED (35,36). We observed lower levels of EMS utilisation (reported in 2 studies; range 1.4-35%), lower bystander CPR rates (reported in 10 studies; range 3-30%), and very low bystander AED rates in the studies included in our review. Variation in response times, and a noteworthy absence of integrated community response systems means that mortality from OHCA in the GCC likely occurs on scene. These findings highlight a significant weakness in the early access and public response to OHCA in the region. Anecdotal reasons for such challenges in the GCC include a lack of defibrillators in public places, “Good Samaritan” laws that stipulate only certified people should provide medical assistance, lack of governing bodies to oversee all aspects of medical emergency response, rurality, and cultural norms (19). Cultural challenges include a lack of knowledge of CPR, a lack of confidence to intervene, and barriers to CPR and AED use, especially on female OHCA patients (37). We also observed over-reliance on private transport rather than EMS to transport OHCA patients, mirroring results in non-OHCA studies in the region (38). Attempts to address such challenges are yielding results, as evidenced in the reported increased use of EMS for OHCA (17,19,20); however, public perceptions related to recognising and intervening in OHCA remain unstudied in the GCC.

Elsewhere internationally, developing integrated community response models has yielded significant improvements in OHCA measures. The Republic of Ireland is a good exemplar, and it shares characteristics of rurality in common with GCC countries (39,40), despite their obvious geographical differences. In 2018 national figures for Ireland (OHCA n=2,442) were reported as: 81% bCPR (excluding EMS witnessed cases), 22% bAED, 26% ROSC and 7.2% (n=176) survival to discharge, the majority with cerebral performance score of 1 or 2 (41). These results represent significant improvements when compared to national figures from only five years prior (n=1,885): 69% bCPR (excluding EMS witnessed cases), 16% bAED, 23% ROSC and 6% survival to discharge (n=120) (42). In that five year period, Ireland further developed a national network of integrated community response schemes, focused on the provision of early CPR, including dispatcher-assisted CPR (DA-CPR). As a result, continued improvements in OHCA measures are likely to be realised in the future. Due to the challenges we observed in this review in relation to access to EMS, bCPR, bAED, and cultural and logistical barriers, we observed low ROSC rates, and subsequent low levels of survival to hospital discharge across the region. In other words, lower survival as an outcome is reflective of process issues in the resuscitation attempt, particularly in the pre-arrival phase.

### Lack of research and coordinated effort

The processes and outcomes related to OHCA are well-researched in a number of countries, in part due to established, coordinated cardiac arrest registries. These may be national [e.g. OHCAR in Ireland (40)] or regional [e.g. EuReCa in Europe (32)], and they allow researchers to examine trends, correlations and concerns related to OHCA with large prospectively-collected datasets. Another example is the Pan-Asian Resuscitation Outcomes Study [PAROS (25)], an international cardiac arrest registry study of which the UAE is a reporting member. Our review also demonstrated that OHCA research efforts are sporadic across the GCC, and even within countries, such research efforts are not strategically coordinated. For example, none of the existing UAE PAROS-member studies reported on OHCA in the Emirate of Abu Dhabi, and thus these publications do not present a complete account of OHCA in the UAE. The heterogeneity of populations within PAROS across Asia and the Middle East presents a challenge when we attempt to utilise it as a solution for the GCC (34). In some GCC countries, OHCA outcomes remain unknown due to a complete lack of reporting. Despite a previous call for a regional initiative for data collection and sharing (43), there has been no concerted effort to create OHCA reporting databases in the GCC. As a result of this lack of coordinated effort, we cannot provide scientific answer to research questions, which creates a barrier to evidence-informed policy and decision making in the GCC. Additionally, the provision of public education, coordination of public information campaigns, bystander and community response to OHCA, strategic planning of emergency medical response, and the coordination of research efforts are (where they exist) spread across multiple local or national agencies within each country. This may lead to a lack of coordinated effort in relation to the management of OHCA. Without such strategic oversight, there is potential for duplication of effort, missed opportunities, and lack of compatibility, both within and between countries in the region.

### Recommendations

The low OHCA survival rates and characteristics observed in our review highlight the regional challenges we face when we attempt to implement existing OHCA management strategies produced by non-GCC systems. Regional challenges include pre-existing morbidity of patients, immigrant populations, unclear “Good Samaritan” laws, low levels of EMS utilisation, lack of AED response, low bystander CPR rates, cultural challenges, and an unknown OHCA recognition rate. As a result, the traditional cardiac arrest ‘chain of survival’ faces challenges as a ‘one size-fits all’ strategy within the region due to such differences in context. Therefore countries within the GCC region may need to customise their implementation of accepted OHCA management strategies to reflect cultural, logistical and demographical variations. To inform such approaches, it is first necessary to highlight regional OHCA characteristics, identify potential OHCA management challenges, and subsequently propose areas for consideration and improvement. Based on our review findings, we provide several recommendations to reduce the OHCA burden in the region (see Textbox 1), largely reflective of the recommendations of the Global Resuscitation Alliance (44). In the first instance, and essential in order to implement additional recommendations, we recommend that a lead agency should be established within each country to coordinate and manage OHCA response within the community. This agency should advocate for the passing of “Good Samaritan Law” equivalents within the region to encourage and protect community responders and bystanders, and may provide leadership on the integration of trained responder dispatch systems (45,46). This agency would continue to develop EMS response, in particular through the delivery of basic life support training, the creation of integrated community response models, and the provision of DA-CPR to improve OHCA process and outcome measures. Community response models empower the local community, and may work to alleviate cultural barriers and the increased response times associated with rurality. Additionally, advocating for uniform emergency numbers, coordinated dispatch centres, and compatible EMS system designs are essential components of early recognition and response in OHCA (47,48).

Public health initiatives should be developed and implemented (or continued where already present) to reduce risk factors for cardiovascular disease, respiratory conditions, and preventable trauma in paediatrics, including drowning. We also recommend targeted public health interventions aimed at improving public health concerns highlighted in expatriate populations. Additionally, in light of the proportion of OHCA that occurred at home in the region (similar to other studies internationally), the lead agency should implement widespread information campaigns through various media, that inform the public on the existence of “Good Samaritan Law” equivalents, the importance of early OHCA recognition, how to access EMS, and the importance of early CPR (which can be further supported via provision of DA-CPR). Furthermore, the lead agency should promote improved access to AEDs in public places that have high incidence of OHCA (49), develop and maintain a national registry of publicly accessible AEDs (50) and reinforce the importance of basic life support training for members of the public. This focus on improved care **prior** to EMS arrival is crucial in improving OHCA outcomes in GCC countries (35). Finally, in order to improve our understanding of OHCA in the region, we advocate for the creation of a reporting database within each GCC country, and a further cooperative database within the region. The lead agency for each country should oversee the development of such a registry, which will enable continued research into the characteristics and management of OHCA in the region. Such databases should align with international data collection and reporting conventions to facilitate data sharing and collaboration (51).

##### Textbox 1. Summary of recommendations.

1. Create a lead agency in each country for strategic oversight of OHCA preparedness, response, management and research
2. Develop community response models, including dispatcher-assisted CPR, and community responder dispatch models
3. Advocate for the legislation of ‘Good Samaritan Law’ equivalents
4. Implement (or continue) public health initiatives to reduce risk factors, including cardiac related health checks for immigrant populations
5. Coordinate widespread public information campaigns related to recognition and management of OHCA, early EMS access, and early CPR
6. Improve access to AEDs in high-risk locations, and create national AED registries
7. Establish and maintain OHCA registries in each country and the GCC region which align with international conventions

### Limitations

Our findings need to be interpreted in the context of certain limitations. We may not have identified all relevant studies despite our attempts to be comprehensive. Our search and review was restricted to articles published in English and Arabic, but this does not inherently bias a review (52). The Google Scholar searches were limited to the first 300 results which is considered adequate for grey literature searches (53). Further grey literature sources may exist that we did not identify; however, we attempted to minimize this risk by using the Grey-Matters Checklist (11), contacting SMEs in the region, and searching non-indexed sources. A number of studies included data on presumed cardiac aetiology as well as traumatic OHCA. We attempted to identify and report only non-traumatic OHCA data where possible, but this was not always possible. We noted in our data where we were able to perform this. Finally, there are a limited number of POHCA studies in our review. Despite these limitations, we propose that our review offers the first comprehensive overview of OHCA in the GCC region.

## Conclusion

Our review is the first to collate and comprehensively report data on OHCA in the GCC region. We highlighted unique demographic traits, and challenges related to OHCA processes and outcomes in the region. We also acknowledged where countries have implemented strategies to address OHCA, and process and outcome measures are improving. Finally, despite the significant burden associated with OHCA in the region, we observed a concerning lack of research and a lack of coordinated effort. In order to address these concerns, we recommend the creation of lead agencies responsible for coordinating and developing OHCA strategies. Such strategies may include implementing community response models including dispatcher-assisted CPR, advocating for “Good Samaritan Law” equivalents, coordinating mass public education campaigns targeted at cultural and logistical issues, supporting public health risk-reduction initiatives, and establishing OHCA reporting databases.

## Data Availability

Contained in Appendices

## Author Declarations

The authors declare that this work has not been published elsewhere. Data from this study were presented at the Emirates Society of Emergency Medicine scientific meeting on December 13 th 2019 in Abu Dhabi, UAE. The authors declare that they are responsible and accountable for the accuracy and integrity of all aspects of this work.

## Author Contributions

AB, CL, SD, SK, MA, SF and FC designed the study. MA and AB designed and conducted the literature searches. CL and AB performed screening and data abstraction. DA and AA contributed additional information to data analysis. AB authored the first draft of the manuscript. All authors contributed to the authoring and editing of the manuscript. All authors have approved the final version for publication.

## Conflicts of interest None declared

Out-of-hospital cardiac arrests in the Gulf Region: a scoping review

